# Vaccines provide disproportional protection to the increased hospitalisation risk posed by the Delta variant of SARS-CoV2: a meta-analysis

**DOI:** 10.1101/2021.12.15.21267799

**Authors:** Mirre J P Simons

**Affiliations:** School of Biosciences, University of Sheffield, Sheffield, United Kingdom. Western Bank, S10 2TN, Sheffield, United Kingdom

## Abstract

Variants of SARS-CoV2 that achieved global dominance (Alpha and Delta) have been associated with increased hospitalisation risk. A quantification of this risk across studies is currently lacking for Delta. Furthermore, how risk for severe disease changes in both vaccinated and unvaccinated individuals is important as the underlying risks determine public health impact. The surplus risk of Delta versus Alpha on hospitalisation was determined using random-effects meta-analysis. Infection with the Delta compared to the Alpha variant increased hospitalisation risk (unvaccinated: log HR 0.62, CI: 0.41 – 0.84, P < 0.0001; linear HR 1.87). This finding should inform our response to future variants of concern, currently Omicron. SARS-CoV2 variants that achieve dominance, have achieved this through a higher rate of infection and this evolutionary trajectory has also come with a correlated higher risk of severe disease. The surplus risk posed by Delta was significantly lower however in the vaccinated (model estimate -0.40, CI: -0.73 – -0.07, P = 0.017). Vaccination thus provided a disproportionate level of protection to hospitalisation with the Delta variant and provides further rationale for vaccination for SARS-CoV2 as a durable public health measure.

## Main text

During the SARS-CoV2 pandemic a key question was do vaccines work? After the emergence of variants focus shifted to whether vaccines still provide protection. Both these questions have been answered with a resounding yes thus far^1^. However, a largely overlooked and understudied question has been if vaccines protect against the increased severe disease risk that has now been suggested for the two variants that achieved global dominance, Alpha (B.1.1.7)^2–4^ and Delta (B 1.617)^5^. A quantification of this risk and its interplay with the protection offered by vaccination across studies is currently lacking for Delta. Vaccines could provide proportional protection, i.e. lower the increased hazard of hospitalisation posed by a variant by a similar proportional amount, could be escaped or provide disproportional protection. Note, such an analysis is different to studying vaccine effectiveness, as this is a ratio of risks resulting from infection with the same variant. The consideration of how risk changes with a new variant is of utmost concern. Even when vaccine effectiveness is unaffected, the underlying risks for both categories, vaccinated and unvaccinated can still be higher (or lower), changing the impact on public health of a variant. The recent emergence of Omicron has again highlighted this importance.

Hospitalisation risk was used as a proxy for disease severity and a meta-analysis was performed across studies to quantify the risk of the Alpha versus the Delta variant in the unvaccinated and vaccinated. The number of studies that estimated the hospitalisation risk of Delta versus Alpha for both vaccinated and unvaccinated individuals is relatively low. Perhaps explained by the demand put on the data to conduct such analyses. Longitudinal data on patients and their outcomes are required and only during the rapid gain in dominance of Delta can reliable comparisons be made. Moreover, adjustment for differences in confounding variables, such as age, are required for effective estimation of the true effect^4^. Five studies^5–9^ included estimates for both vaccinated and unvaccinated individuals. Three studies^10–12^ were identified with known vaccination status from which the hospitalisation risk of Delta could be extracted only for the unvaccinated. Vaccination was not separated by vaccine type in these studies but given the countries where the included studies were conducted in, the vaccines used are a combination of BNT162b2, mRNA-1273 and ChAdOx1 nCoV-19.

Infection with the Delta compared to the Alpha variant increased hospitalisation risk (Figure 1; Table S1; unvaccinated: log HR 0.62, CI: 0.41 – 0.84, P < 0.0001; linear HR 1.87). This risk was significantly lower in the vaccinated individuals (Table S1, model estimate of vaccination: log HR -0.40, CI: -0.73 – -0.07, P = 0.017). When data was split for first and second doses, second doses showed the strongest response (Table S2, but note only two studies included estimates for single dosage). Estimates of these vaccination status specific risks remained similar and statistically significant when the three studies that only estimated risk in the unvaccinated were excluded (Table S3-S4). Within the vaccinated individuals there was an increased risk posed by Delta (log HR 0.20, CI: -0.1766 – 0.5842; linear HR 1.22) but this was not significantly different from zero (P = 0.29). Vaccination provides a disproportionate level of protection to hospitalisation with the Delta variant.

**Figure 1.**
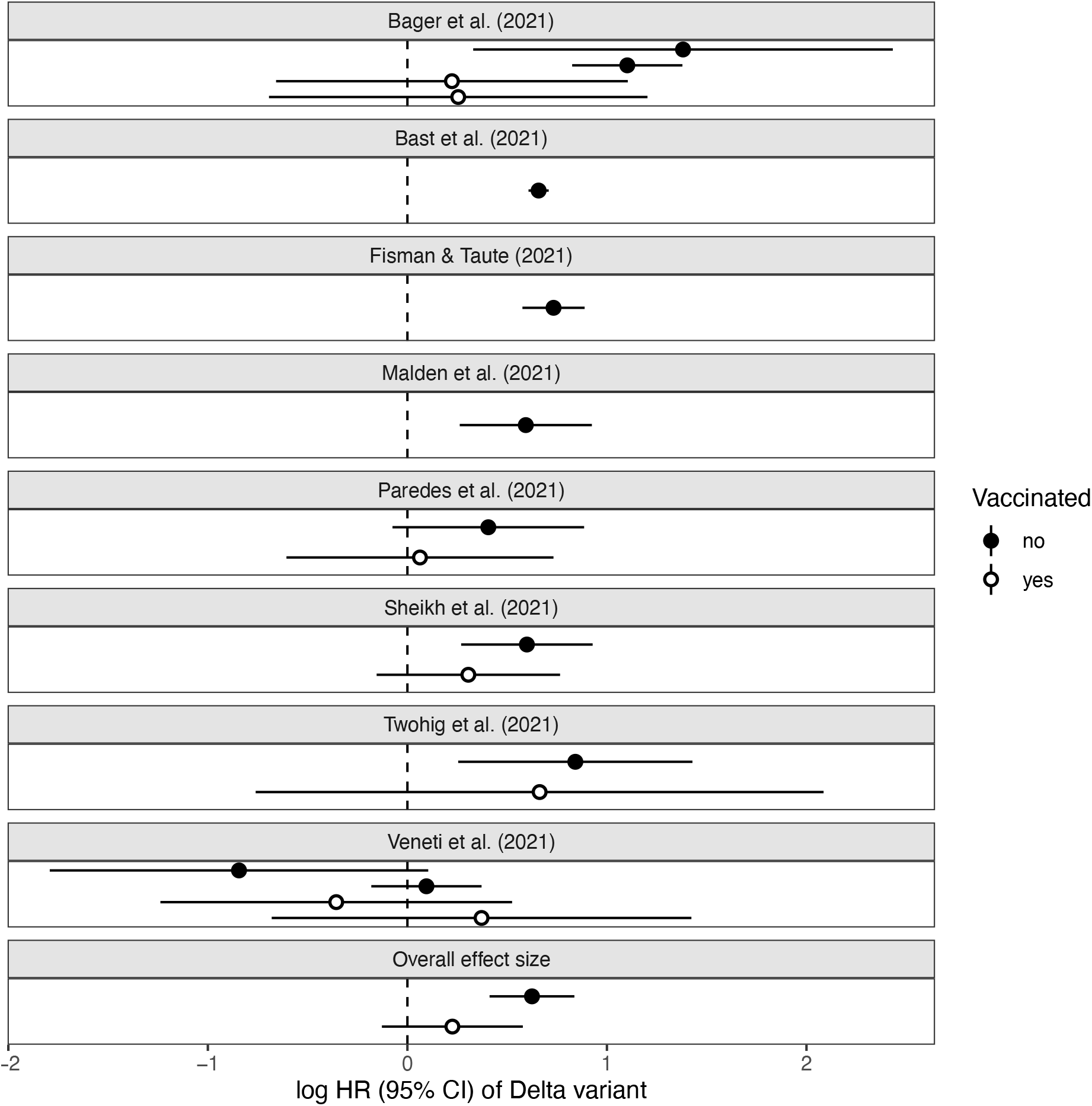
Individual effect sizes per study for hospitalisation risk of Delta versus Alpha, split for vaccination status. Overall effect sizes as estimated through random-effects meta-analysis are shown at the bottom of the figure.

As with the Alpha variant, the Delta variant is associated with a larger risk of severe disease, most prominent in the unvaccinated. This finding should inform our response to future variants of concern, currently Omicron. SARS-CoV2 variants that achieve dominance, have achieved this through a higher rate of infection and this evolutionary trajectory has also come with a correlated higher risk of severe disease. These consequences were now seen twice during the pandemic, with both Alpha and Delta. Fortunately, vaccination reduces hospitalisation risk. For Delta there is even a disproportionate rescue of surplus hospitalisation risk compared to Alpha (Figure 1), and limited data available for Alpha also support this notion compared to the wildtype variant (two studies^7,13^, Table S5). Such a response could be due to several currently unknown reasons. For example, specific aspects of immunity^14^ primed by vaccination or a disproportionate rescue by vaccination of the gained virulence of variants of concern.

The data summarised here through meta-analysis provide unique insight into how variants of concern have impacted the SARS-CoV2 pandemic. The disproportionate rescue from hospitalisation risk through vaccination identified provides additional support for vaccination as a durable public health intervention. The gain in virulence in both Alpha and Delta in the unvaccinated does pose a warning for future variants when vaccination coverage is low or spread of the variant is wide. In practice, estimating the impact of novel variants of concern on risk to develop severe disease can only be conducted on a very small timescale and easily becomes confounded^4^ by e.g. improvements in treatment, changes in demography and vaccine waning^15^. With both Alpha, and Delta, and now Omicron, a loss (or regain) of template amplification in the majority of qPCR tests provided a reliable and convenient proxy for which variant a patient is infected with. Future variants need not provide such convenience. Moreover, the faster a variant gains dominance the smaller the time window in which accurate comparisons can be made. In that case, in a public health setting a rapid shift in the demography (e.g. vaccinated versus unvaccinated) and number of hospitalisations over time is probably most informative of the severity of a variant of concern identified and its interaction with vaccination status. The prior known history of evolutionarily successful variants showing increased severe disease risk would however warrant caution upon identification of spread of a novel variant over relying on real-time data.

## Methods

Literature search (final search 10 December 2021) was conducted in Google Scholar using the following search terms: “hospitalisation OR hospitalization” risk B.1.1.7 B 1.617 vaccinated (“hazard ratio” OR HR). Inclusion criteria were that hospitalisation risk of Delta versus Alpha infection was reported per vaccination category. Hospitalisation risk did not include emergency room attendance if this was reported separately. Adjusted hazard (or risk) ratios were extracted as adjusted for the covariates included in each study. No exclusion criteria were formulated. The search yielded 230 results (see PRISMA guide, Figure S1) and returned 8 eligible studies. For the post-hoc search for Alpha specific papers (yielding two studies), the literature was queried again and references within papers collated for the Delta search selecting for studies that included estimates for both vaccinated and unvaccinated individuals. We collected adjusted hazard or risk ratios as reported in text, tables or derived from combining coefficients from models reported. These were log (natural) transformed and the corresponding standard errors were derived from the reported linear confidence intervals for use in meta-analysis. Random effects meta-analysis was conducted in *metafor*^16^ in R and included study as a random term, to correct for multiple effect sizes per study.

## Data Availability

All data is presented in the manuscript. All R scripts available from author.

## Supplementary Tables

**Table S1.**
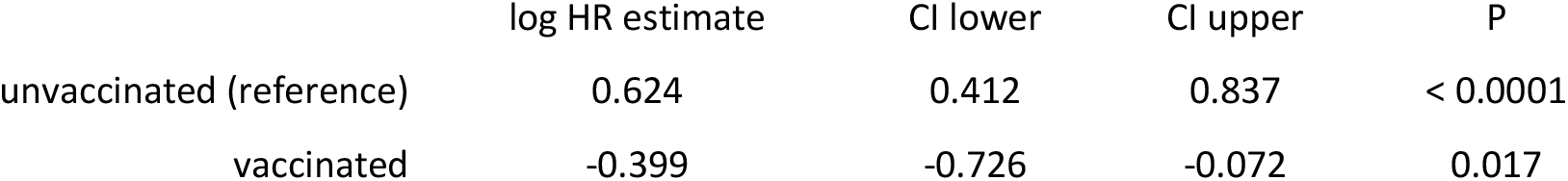
Meta-analytic model estimating the risk of Delta versus Alpha for hospitalisation across the 8 studies included.

|  | log HR estimate | CI lower | CI upper | P |
| --- | --- | --- | --- | --- |
| unvaccinated (reference) | 0.624 | 0.412 | 0.837 | < 0.0001 |
| vaccinated | -0.399 | -0.726 | -0.072 | 0.017 |

**Table S2.** Meta-analytic model estimating the risk of Delta versus Alpha for hospitalisation across the 8 studies included, separated by vaccination dose. Note only two estimates for single dosing were available across the dataset.

|  | log HR estimate | CI lower | CI upper | P |
| --- | --- | --- | --- | --- |
| unvaccinated (reference) | 0.625 | 0.413 | 0.838 | < 0.0001 |
| one dose | -0.300 | -1.025 | 0.424 | 0.417 |
| two doses | -0.421 | -0.780 | -0.063 | 0.021 |

**Table S3.** Meta-analytic model estimating the risk of Delta versus Alpha for hospitalisation across the 5 studies included that estimated risk within vaccinated and unvaccinated individuals.

|  | log HR estimate | CI lower | CI upper | P |
| --- | --- | --- | --- | --- |
| unvaccinated (reference) | 0.596 | 0.231 | 0.961 | 0.001 |
| vaccinated | -0.388 | -0.721 | -0.056 | 0.022 |

**Table S4.** Meta-analytic model estimating the risk of Delta versus Alpha for hospitalisation across the 5 studies included that estimated risk within the vaccinated and unvaccinated individuals, separated by vaccination dose. Note only two estimates for single dosing were available across the dataset.

|  | log HR estimate | CI lower | CI upper | P |
| --- | --- | --- | --- | --- |
| unvaccinated (reference) | 0.598 | 0.232 | 0.963 | 0.001 |
| one dose | -0.295 | -1.021 | 0.432 | 0.427 |
| two doses | -0.410 | -0.775 | -0.045 | 0.028 |

**Table S5.** Meta-analytic model estimating the risk of Delta versus Alpha for hospitalisation across the 2 studies that estimated the risk of Alpha compared to Wildtype variant in vaccinated and unvaccinated individuals.

|  | log HR estimate | CI lower | CI upper | P |
| --- | --- | --- | --- | --- |
| unvaccinated (reference) | 0.360 | 0.229 | 0.490 | < 0.0001 |
| vaccinated | -0.403 | -0.895 | 0.089 | 0.108 |

---

**Supplementary Figure 1.**
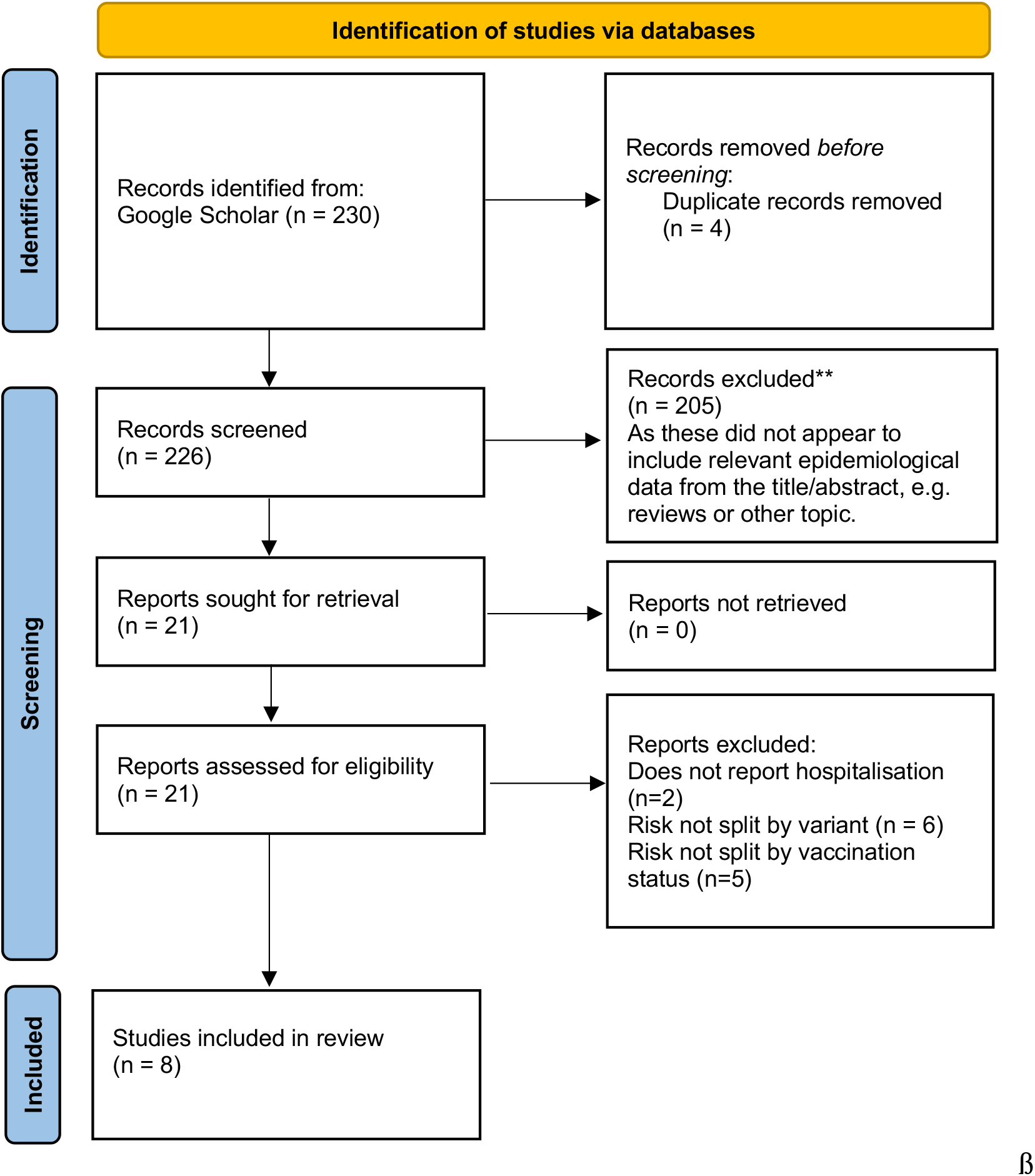
PRISMA Flow diagram of the literature search conducted.

